## Supplemental material for "Dengue severity by serotype and immune status in 19 years of pediatric clinical studies in Nicaragua"

**This pdf file includes:**

Table S1

Figures S1 to S9

**Supplementary Table S1. Classification of dengue disease severity by 1997 and 2009 World Health Organization criteria and definition of clinical variables.**

| | |
|---|---|
| **Case definition** | |
| Suspected dengue | Fever with 2 or more of the following: nausea or vomiting; headache, retroorbital pain, myalgia, arthralgia, leukopenia (WBC ≤4000 cells/$mm^3$), rash, positive tourniquet test; any warning sign (PDHS only) |
| **Clinical variables** | |
| Fever | >37.5ºC |
| Hemoconcentration | ≥20% increase in hematocrit (compared to the stabilized hematocrit at hospital discharge) |
| Hypotension | Systolic blood pressure <80 mmHg for children <5 years of age and <90 mmHg for children ≥5 years of age |
| Leukopenia | WBC ≤4000 cells/mm3 |
| Narrow pulse pressure | Difference between systolic and diastolic blood pressure ≤20 mmHg. |
| Plasma leakage | Any of the following:<br>- Fluid accumulation: pleural effusion, ascites/free fluid in abdomen, pericarditis, gallbladder wall thickening >3mm (by ultrasound)<br>- Hematocrit change ≥20% during illness<br>- Low albumin <3.5 |
| Poor capillary refill | >2 sec |
| Pleural effusion | Clinically, through ultrasound, or X-ray, with the latter being the most sensitive. Clinically, there is a decrease in breathing sounds. X-ray shows evidence of fluid on radiography. Excessive accumulation of fluid in the pleural space (normal volume is 15 to 20 ml). When it exceeds 30 ml, it can be diagnosed by ultrasound. |
| Ascites | Presence of fluid in the abdominal cavity, either through clinical examination or ultrasound. |
| Mucosal bleeding | Gingivorrhagia, epitaxis, subconjunctival bleeding, hematemesis, hemoptisis, melena, vaginal bleeding, or hematuria. |
| Persistent vomiting | ≥3 episodes in 12 hour period |
| Respiratory distress | Tachypnea and any of the following: oxygen saturation les 92, accessory muscle use, nasal flaring, altered gasometrical parameters |
| **1997 WHO classification** | |
| Dengue fever (DF) | Fever with 2 or more of the following: headache; retro-orbital pain; myalgia; arthralgia; leukopenia (WBC ≤4000 cells/mm3); rash; haemorrhagic manifestations<br>AND not fulfilling criteria for DHF or DSS |

| | |
|---|---|
| Dengue hemorrhagic fever (DHF) | All of the following must be present:<br>- Fever or history of acute fever lasting 2–7 days<br>- Haemorrhagic manifestations (positive tourniquet test; petechiae, equimosis, purpura or bleeding from mucosa, gastrointestinal tract, injection sites or other locations; hematemesis; melena)<br>- Thrombocytopenia (≤100,000 platelets/mm3)<br>- Evidence of plasma leakage due to increased vascular permeability |
| Dengue shock syndrome (DSS) | DHF with hypotension for age or narrow pulse pressure (≤20 mmHg) plus one of the following: rapid and weak pulse; cold, clammy skin; restlessness; poor capillary refill (>2 sec) |
| DHF/DSS | DHF or DSS |
| **2009 WHO criteria** | |
| Dengue without warning signs (DwoWS) | Confirmed dengue without any warning sign |
| Dengue with warning signs (DwWS) | Persistent vomiting, abdominal pain, mucosal hemorrhagic manifestations, plasma leakage, hepatomegaly, irritability, , platelet count ≤100,000 cells/mm$^3$, increase of haematocrit with decrease of 10,000 platelets / mm$^3$ in 24 hours or increase of haematocrit with decrease of platelet count ≤100,000 cells/mm$^3$ any time period |
| Severe dengue (SD) | (1)Severe plasma leakage leading to: shock or respiratory distress OR<br>(2) severe bleeding; OR<br>(3) severe organ impairment |
| (1a) Severe plasma leakage (plasma leakage and respiratory compromise) | 1. Respiratory distress, AND<br>1. 2. shock |
| (1b) shock<br>hypotensive Shock<br><br>Compensated shock | Narrow pulse pressure ≤20 mmHg, hypotension for age and sex and any of the following; capillary refill >2 seg, elevated heart rate, elevated breathing rate, , weak pulse, pale and/or cold skin.<br><br>capillary refill >2 seg and any of the following: elevated heart rate, elevated breathing rate, , weak pulse, pale and/or cold skin, |
| (2) Severe bleeding | Any bleeding with life threatening condition evaluated by clinician |
| (3) Severe organ involvement | AST or ALT ≥1000 U, intubation/mechanical ventilation, encephalopathy, myocarditis, acute hepatitis, Glasgow score <10 blantyre <5, |
| Transition from compensated to hypotensive | Patients with criteria for compensated shock that evolved into hypotension or shortening of pulse pressure despite medical intervention. |

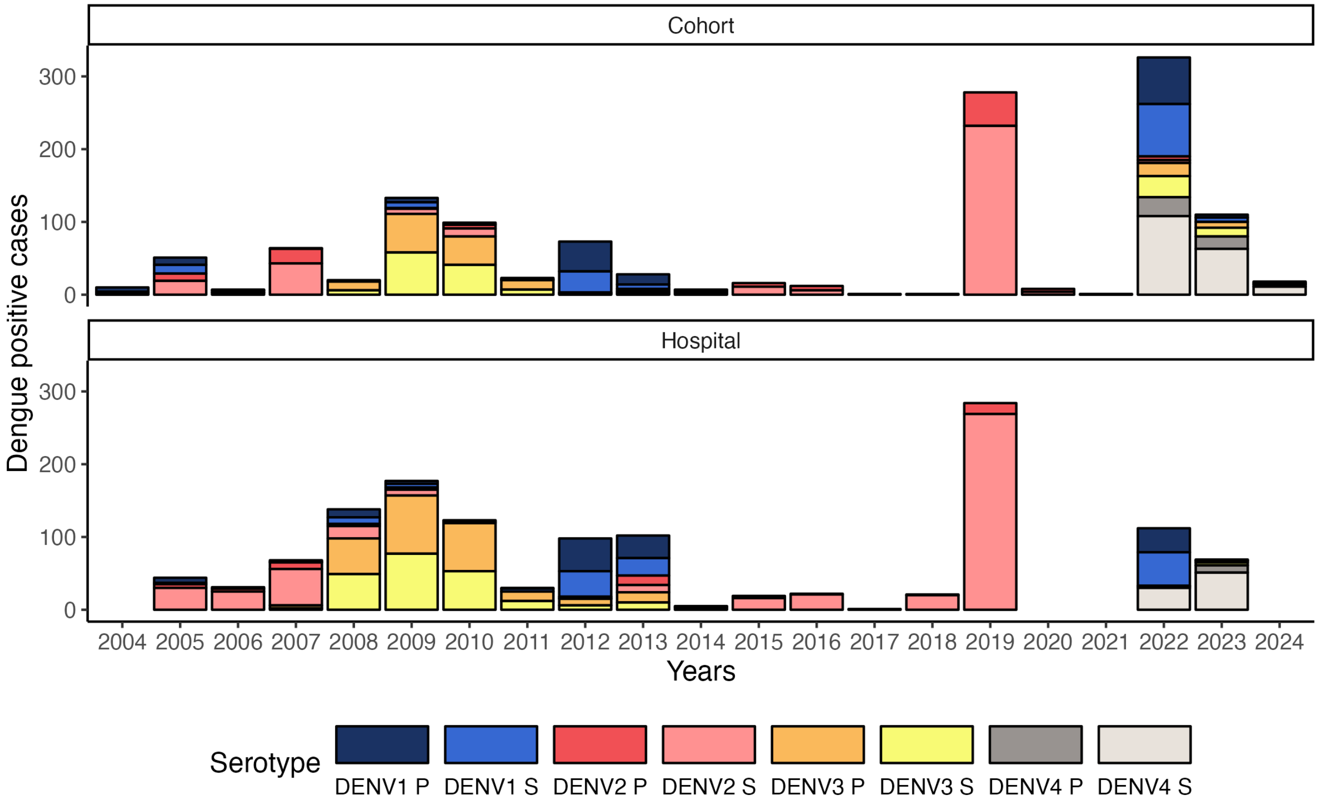


**Supplementary Figure 1. DENV circulation by immune status stratified by study.** Upper panel, Pediatric Dengue Cohort Study; lower panel, Pediatric Hospital-based Study in Managua, Nicaragua, 2004-2022. Circulating serotypes and immune status in Managua are represented. The hospital-based study was not run in 2020 and 2021 due to COVID-19.

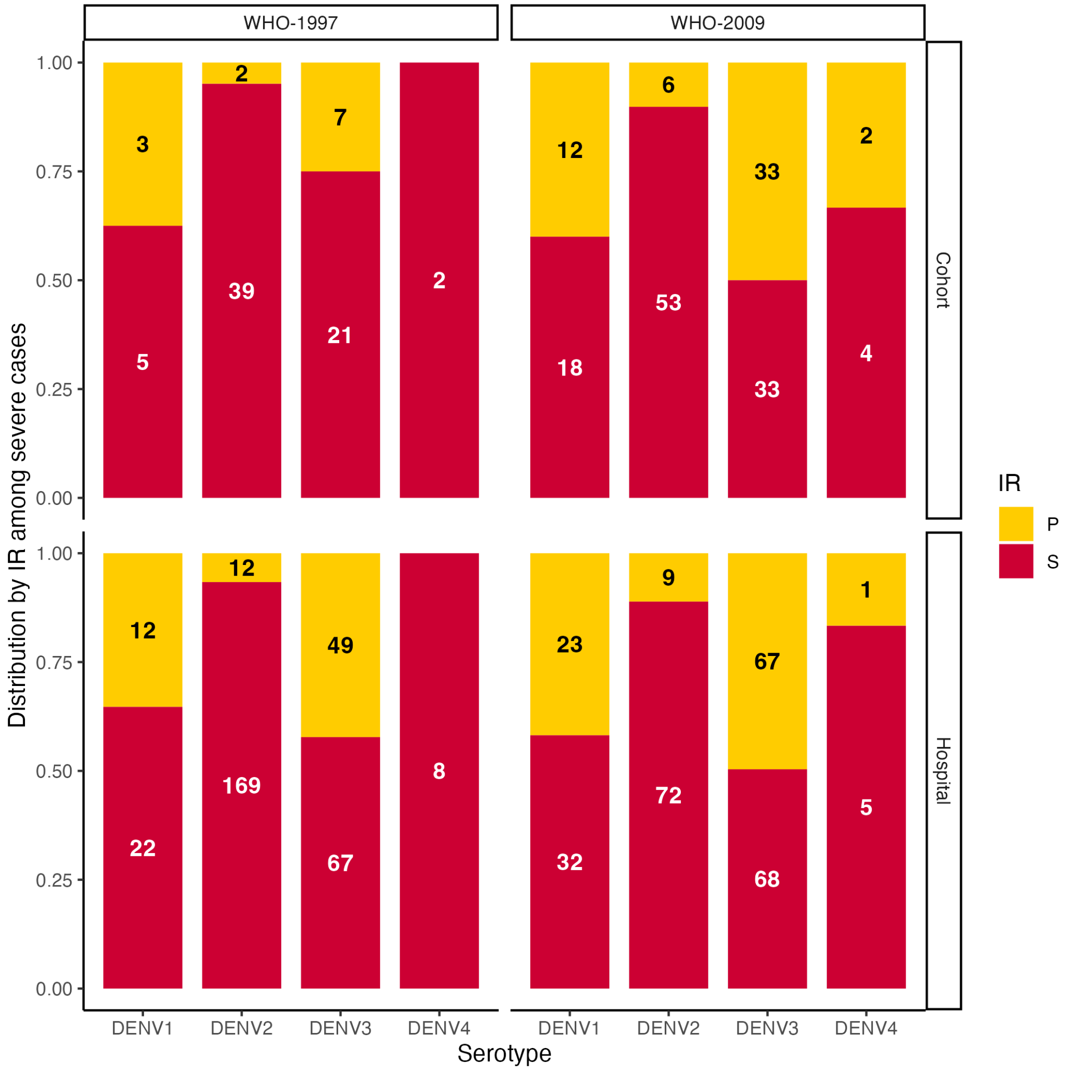


**Supplementary Figure 2. Distribution by immune response among severe dengue cases, stratified by study, Managua, 2004-2022.** Upper panel, Pediatric Dengue Cohort Study (Cohort); lower panel, Pediatric Dengue Hospital-based **S**tudy (Hospital). IR, immune response.

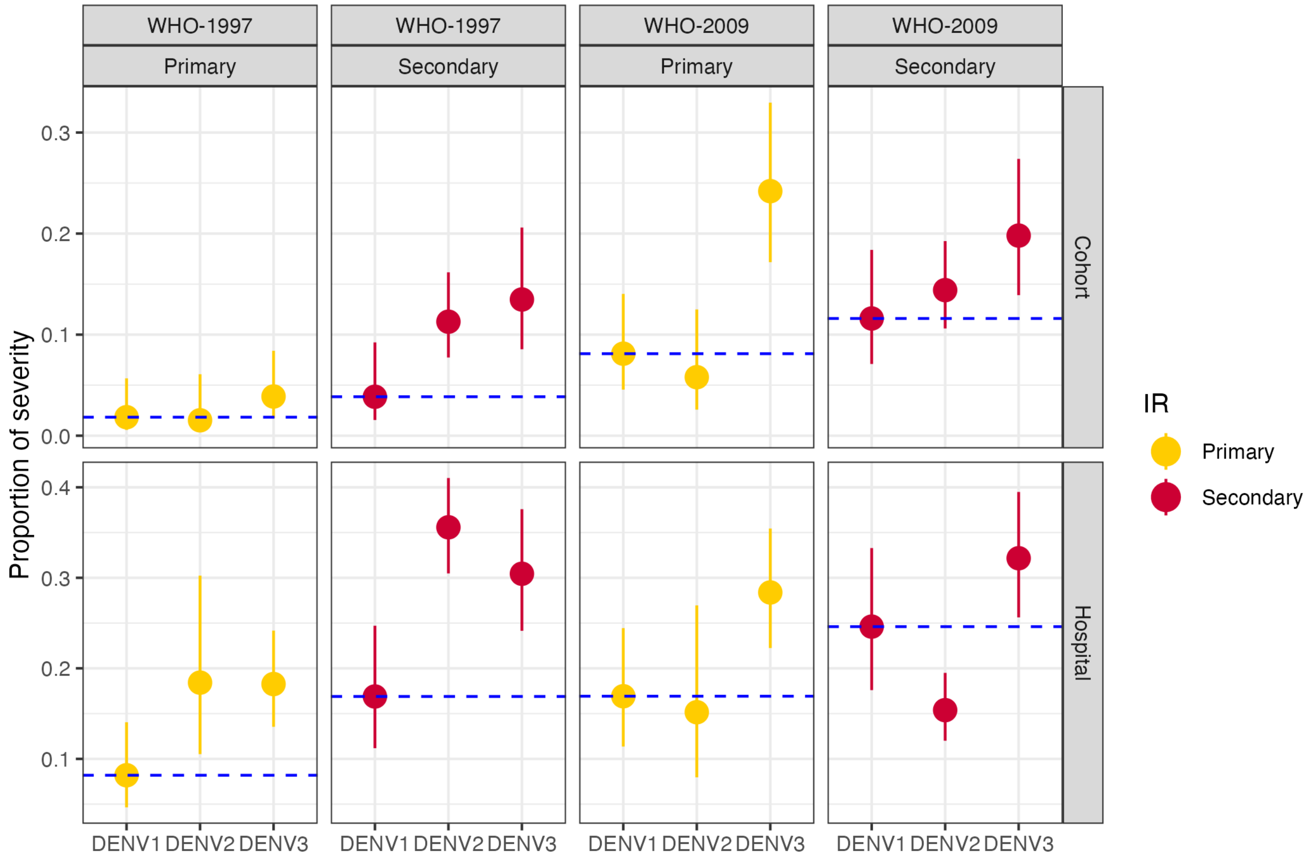


**Supplementary Figure 3. Severity by infecting DENV serotype and immune status by study, Managua, 2004-2022.** Upper panel, Pediatric Dengue Cohort Study (Cohort); lower panel, Pediatric Dengue Hospital-based Study (Hospital). Due to sample size, DENV-4 was excluded from the analysis. IR, immune response.

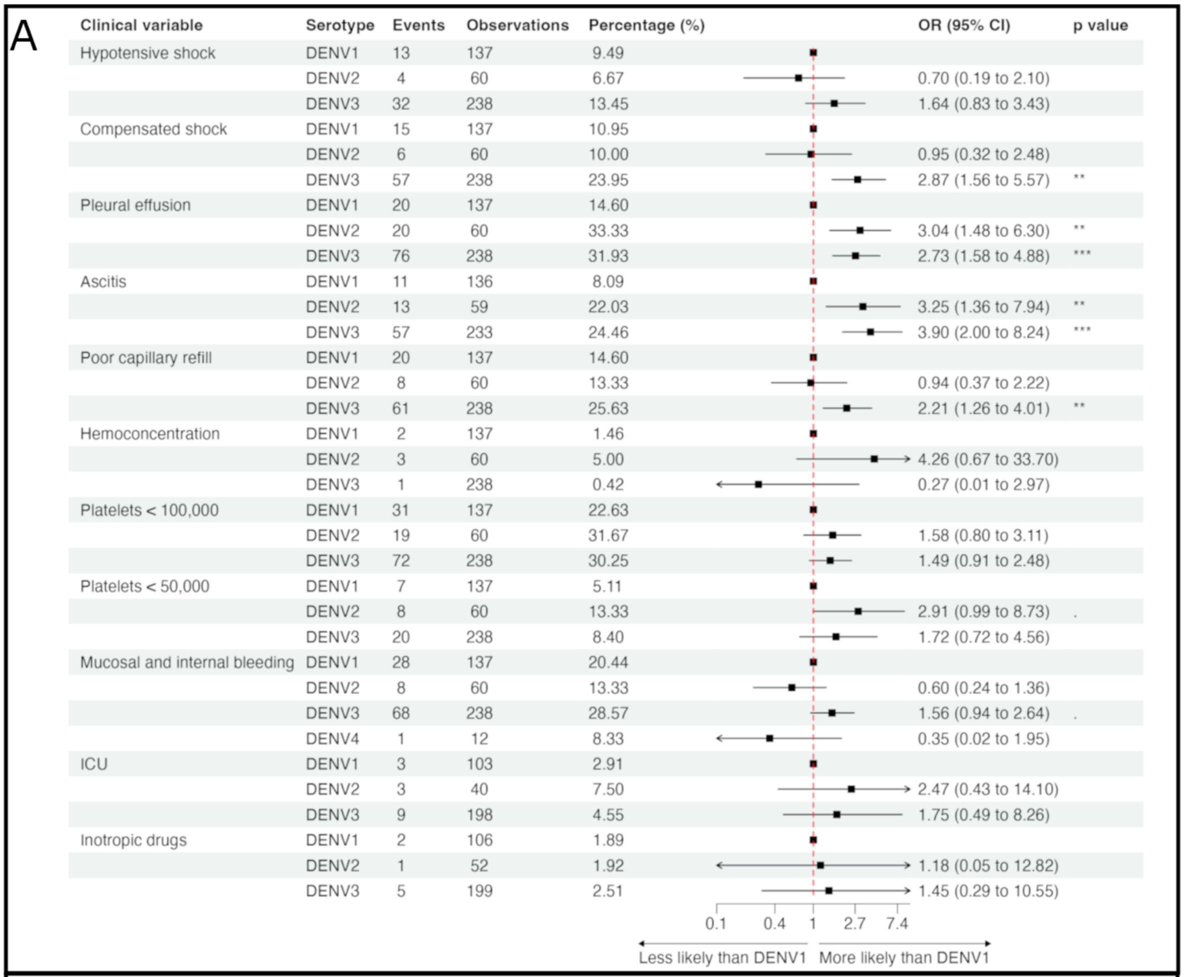

| Clinical variable | Serotype | Events | Observations | Percentage (%) | OR (95% CI) | p value |
|---|---|---|---|---|---|---|
| Hypotensive shock | DENV1 | 13 | 137 | 9.49 | | |
| | DENV2 | 4 | 60 | 6.67 | 0.70 (0.19 to 2.10) | |
| | DENV3 | 32 | 238 | 13.45 | 1.64 (0.83 to 3.43) | |
| Compensated shock | DENV1 | 15 | 137 | 10.95 | | |
| | DENV2 | 6 | 60 | 10.00 | 0.95 (0.32 to 2.48) | |
| | DENV3 | 57 | 238 | 23.95 | 2.87 (1.56 to 5.57) | ** |
| Pleural effusion | DENV1 | 20 | 137 | 14.60 | | |
| | DENV2 | 20 | 60 | 33.33 | 3.04 (1.48 to 6.30) | ** |
| | DENV3 | 76 | 238 | 31.93 | 2.73 (1.58 to 4.88) | *** |
| Ascitis | DENV1 | 11 | 136 | 8.09 | | |
| | DENV2 | 13 | 59 | 22.03 | 3.25 (1.36 to 7.94) | ** |
| | DENV3 | 57 | 233 | 24.46 | 3.90 (2.00 to 8.24) | *** |
| Poor capillary refill | DENV1 | 20 | 137 | 14.60 | | |
| | DENV2 | 8 | 60 | 13.33 | 0.94 (0.37 to 2.22) | |
| | DENV3 | 61 | 238 | 25.63 | 2.21 (1.26 to 4.01) | ** |
| Hemoconcentration | DENV1 | 2 | 137 | 1.46 | | |
| | DENV2 | 3 | 60 | 5.00 | 4.26 (0.67 to 33.70) | |
| | DENV3 | 1 | 238 | 0.42 | 0.27 (0.01 to 2.97) | |
| Platelets < 100,000 | DENV1 | 31 | 137 | 22.63 | | |
| | DENV2 | 19 | 60 | 31.67 | 1.58 (0.80 to 3.11) | |
| | DENV3 | 72 | 238 | 30.25 | 1.49 (0.91 to 2.48) | |
| Platelets < 50,000 | DENV1 | 7 | 137 | 5.11 | | |
| | DENV2 | 8 | 60 | 13.33 | 2.91 (0.99 to 8.73) | . |
| | DENV3 | 20 | 238 | 8.40 | 1.72 (0.72 to 4.56) | |
| Mucosal and internal bleeding | DENV1 | 28 | 137 | 20.44 | | |
| | DENV2 | 8 | 60 | 13.33 | 0.60 (0.24 to 1.36) | |
| | DENV3 | 68 | 238 | 28.57 | 1.56 (0.94 to 2.64) | . |
| | DENV4 | 1 | 12 | 8.33 | 0.35 (0.02 to 1.95) | |
| ICU | DENV1 | 3 | 103 | 2.91 | | |
| | DENV2 | 3 | 40 | 7.50 | 2.47 (0.43 to 14.10) | |
| | DENV3 | 9 | 198 | 4.55 | 1.75 (0.49 to 8.26) | |
| Inotropic drugs | DENV1 | 2 | 106 | 1.89 | | |
| | DENV2 | 1 | 52 | 1.92 | 1.18 (0.05 to 12.82) | |
| | DENV3 | 5 | 199 | 2.51 | 1.45 (0.29 to 10.55) | |



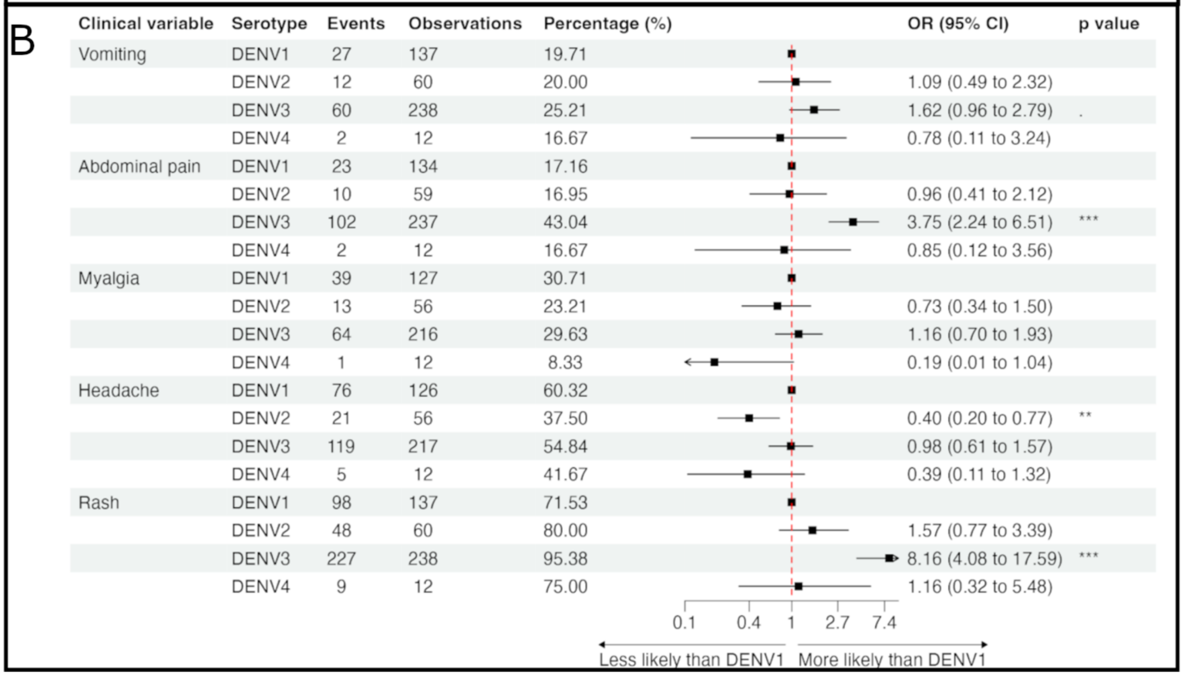

| Clinical variable | Serotype | Events | Observations | Percentage (%) | OR (95% CI) | p value |
|---|---|---|---|---|---|---|
| Vomiting | DENV1 | 27 | 137 | 19.71 | | |
| | DENV2 | 12 | 60 | 20.00 | 1.09 (0.49 to 2.32) | |
| | DENV3 | 60 | 238 | 25.21 | 1.62 (0.96 to 2.79) | . |
| | DENV4 | 2 | 12 | 16.67 | 0.78 (0.11 to 3.24) | |
| Abdominal pain | DENV1 | 23 | 134 | 17.16 | | |
| | DENV2 | 10 | 59 | 16.95 | 0.96 (0.41 to 2.12) | |
| | DENV3 | 102 | 237 | 43.04 | 3.75 (2.24 to 6.51) | *** |
| | DENV4 | 2 | 12 | 16.67 | 0.85 (0.12 to 3.56) | |
| Myalgia | DENV1 | 39 | 127 | 30.71 | | |
| | DENV2 | 13 | 56 | 23.21 | 0.73 (0.34 to 1.50) | |
| | DENV3 | 64 | 216 | 29.63 | 1.16 (0.70 to 1.93) | |
| | DENV4 | 1 | 12 | 8.33 | 0.19 (0.01 to 1.04) | |
| Headache | DENV1 | 76 | 126 | 60.32 | | |
| | DENV2 | 21 | 56 | 37.50 | 0.40 (0.20 to 0.77) | ** |
| | DENV3 | 119 | 217 | 54.84 | 0.98 (0.61 to 1.57) | |
| | DENV4 | 5 | 12 | 41.67 | 0.39 (0.11 to 1.32) | |
| Rash | DENV1 | 98 | 137 | 71.53 | | |
| | DENV2 | 48 | 60 | 80.00 | 1.57 (0.77 to 3.39) | |
| | DENV3 | 227 | 238 | 95.38 | 8.16 (4.08 to 17.59) | *** |
| | DENV4 | 9 | 12 | 75.00 | 1.16 (0.32 to 5.48) | |



**Supplementary Figure 4. Clinical signs and symptoms of dengue in primary cases by serotype in participants of the Pediatric Dengue Hospital-based study, 2004-2022.** A) Clinical signs, laboratory results, and clinical management. B) Symptoms of dengue**.**

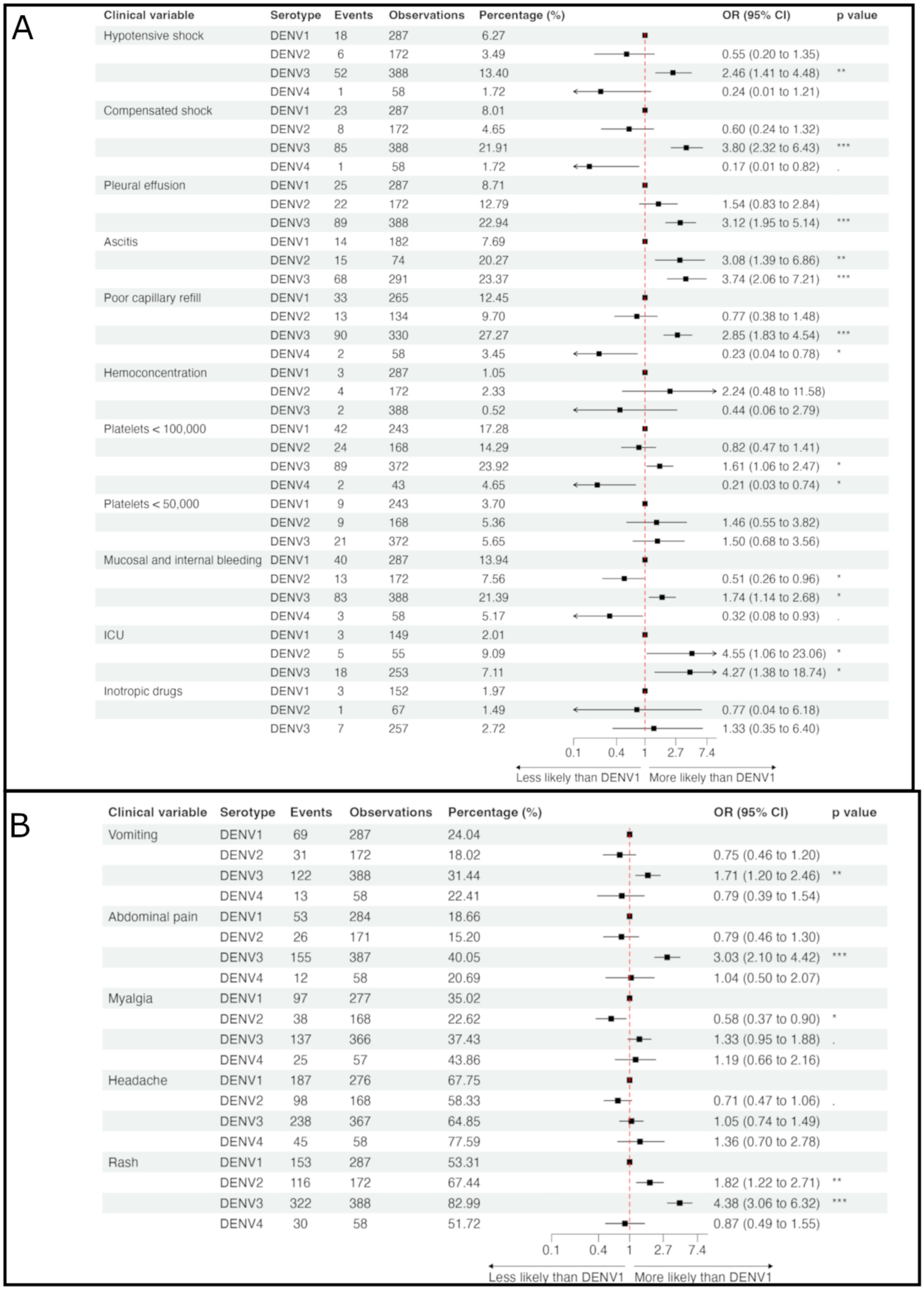


**Supplementary Figure 5. Clinical signs and symptoms of dengue in primary cases by serotype in participants of the Pediatric Dengue Cohort Study transferred to the Pediatric Dengue Hospital-based study, 2004-2022.** A) Clinical signs, laboratory results, and clinical management. B) Symptoms of dengue.

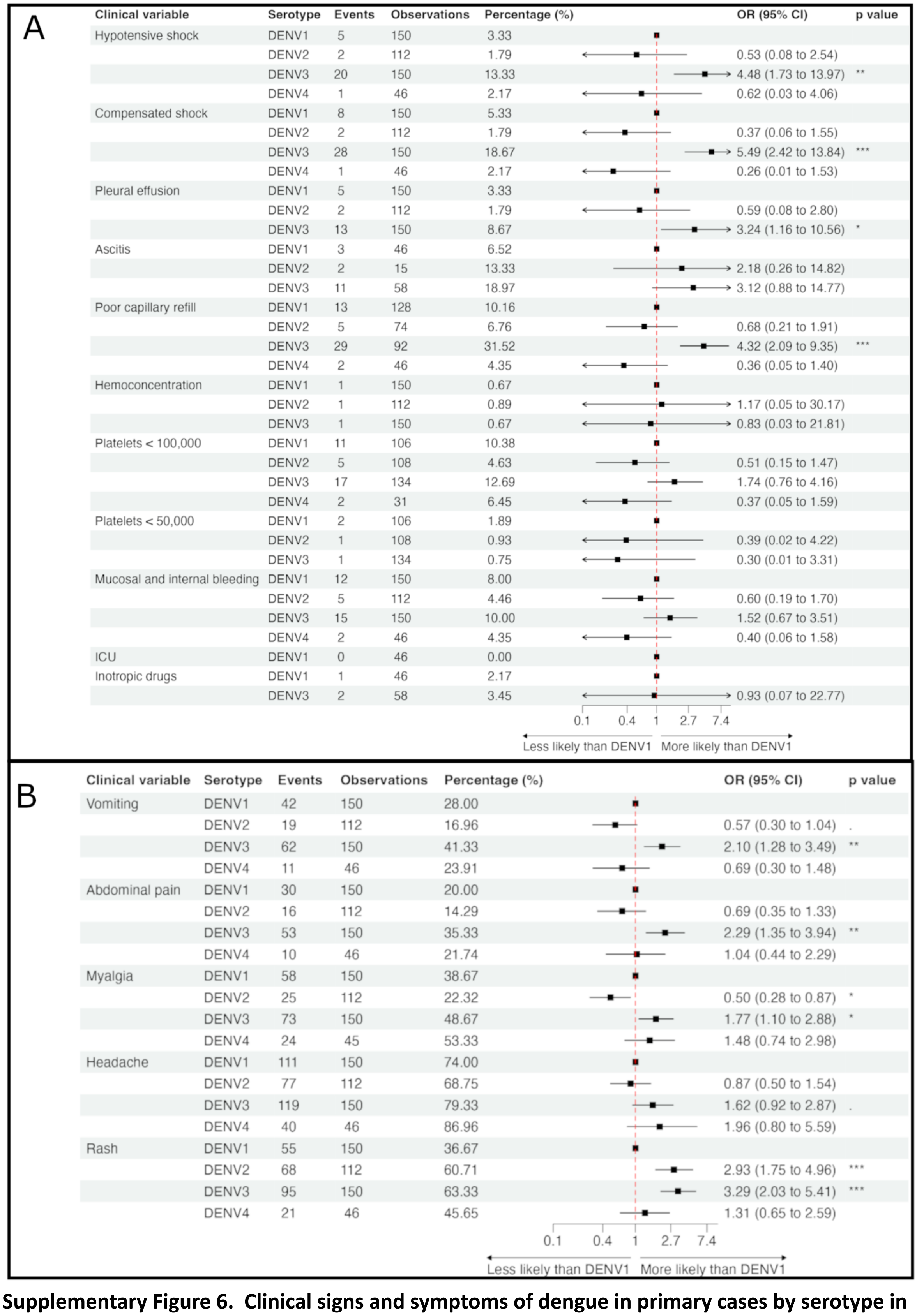

A

| Clinical variable | Serotype | Events | Observations | Percentage (%) | OR (95% CI) | p value |
|---|---|---|---|---|---|---|
| Hypotensive shock | DENV1 | 5 | 150 | 3.33 | | |
| | DENV2 | 2 | 112 | 1.79 | 0.53 (0.08 to 2.54) | |
| | DENV3 | 20 | 150 | 13.33 | 4.48 (1.73 to 13.97) | ** |
| | DENV4 | 1 | 46 | 2.17 | 0.62 (0.03 to 4.06) | |
| Compensated shock | DENV1 | 8 | 150 | 5.33 | | |
| | DENV2 | 2 | 112 | 1.79 | 0.37 (0.06 to 1.55) | |
| | DENV3 | 28 | 150 | 18.67 | 5.49 (2.42 to 13.84) | *** |
| | DENV4 | 1 | 46 | 2.17 | 0.26 (0.01 to 1.53) | |
| Pleural effusion | DENV1 | 5 | 150 | 3.33 | | |
| | DENV2 | 2 | 112 | 1.79 | 0.59 (0.08 to 2.80) | |
| | DENV3 | 13 | 150 | 8.67 | 3.24 (1.16 to 10.56) | * |
| Ascitis | DENV1 | 3 | 46 | 6.52 | | |
| | DENV2 | 2 | 15 | 13.33 | 2.18 (0.26 to 14.82) | |
| | DENV3 | 11 | 58 | 18.97 | 3.12 (0.88 to 14.77) | |
| Poor capillary refill | DENV1 | 13 | 128 | 10.16 | | |
| | DENV2 | 5 | 74 | 6.76 | 0.68 (0.21 to 1.91) | |
| | DENV3 | 29 | 92 | 31.52 | 4.32 (2.09 to 9.35) | *** |
| | DENV4 | 2 | 46 | 4.35 | 0.36 (0.05 to 1.40) | |
| Hemoconcentration | DENV1 | 1 | 150 | 0.67 | | |
| | DENV2 | 1 | 112 | 0.89 | 1.17 (0.05 to 30.17) | |
| | DENV3 | 1 | 150 | 0.67 | 0.83 (0.03 to 21.81) | |
| Platelets < 100,000 | DENV1 | 11 | 106 | 10.38 | | |
| | DENV2 | 5 | 108 | 4.63 | 0.51 (0.15 to 1.47) | |
| | DENV3 | 17 | 134 | 12.69 | 1.74 (0.76 to 4.16) | |
| | DENV4 | 2 | 31 | 6.45 | 0.37 (0.05 to 1.59) | |
| Platelets < 50,000 | DENV1 | 2 | 106 | 1.89 | | |
| | DENV2 | 1 | 108 | 0.93 | 0.39 (0.02 to 4.22) | |
| | DENV3 | 1 | 134 | 0.75 | 0.30 (0.01 to 3.31) | |
| Mucosal and internal bleeding | DENV1 | 12 | 150 | 8.00 | | |
| | DENV2 | 5 | 112 | 4.46 | 0.60 (0.19 to 1.70) | |
| | DENV3 | 15 | 150 | 10.00 | 1.52 (0.67 to 3.51) | |
| | DENV4 | 2 | 46 | 4.35 | 0.40 (0.06 to 1.58) | |
| ICU | DENV1 | 0 | 46 | 0.00 | | |
| Inotropic drugs | DENV1 | 1 | 46 | 2.17 | | |
| | DENV3 | 2 | 58 | 3.45 | 0.93 (0.07 to 22.77) | |



B

| Clinical variable | Serotype | Events | Observations | Percentage (%) | OR (95% CI) | p value |
|---|---|---|---|---|---|---|
| Vomiting | DENV1 | 42 | 150 | 28.00 | | |
| | DENV2 | 19 | 112 | 16.96 | 0.57 (0.30 to 1.04) | . |
| | DENV3 | 62 | 150 | 41.33 | 2.10 (1.28 to 3.49) | ** |
| | DENV4 | 11 | 46 | 23.91 | 0.69 (0.30 to 1.48) | |
| Abdominal pain | DENV1 | 30 | 150 | 20.00 | | |
| | DENV2 | 16 | 112 | 14.29 | 0.69 (0.35 to 1.33) | |
| | DENV3 | 53 | 150 | 35.33 | 2.29 (1.35 to 3.94) | ** |
| | DENV4 | 10 | 46 | 21.74 | 1.04 (0.44 to 2.29) | |
| Myalgia | DENV1 | 58 | 150 | 38.67 | | |
| | DENV2 | 25 | 112 | 22.32 | 0.50 (0.28 to 0.87) | * |
| | DENV3 | 73 | 150 | 48.67 | 1.77 (1.10 to 2.88) | * |
| | DENV4 | 24 | 45 | 53.33 | 1.48 (0.74 to 2.98) | |
| Headache | DENV1 | 111 | 150 | 74.00 | | |
| | DENV2 | 77 | 112 | 68.75 | 0.87 (0.50 to 1.54) | |
| | DENV3 | 119 | 150 | 79.33 | 1.62 (0.92 to 2.87) | . |
| | DENV4 | 40 | 46 | 86.96 | 1.96 (0.80 to 5.59) | |
| Rash | DENV1 | 55 | 150 | 36.67 | | |
| | DENV2 | 68 | 112 | 60.71 | 2.93 (1.75 to 4.96) | *** |
| | DENV3 | 95 | 150 | 63.33 | 3.29 (2.03 to 5.41) | *** |
| | DENV4 | 21 | 46 | 45.65 | 1.31 (0.65 to 2.59) | |



**Supplementary Figure 6. Clinical signs and symptoms of dengue in primary cases by serotype in participants of the Pediatric Dengue Cohort Study, 2004-2022.** A) Clinical signs, laboratory results, and clinical management. B) Symptoms of dengue.

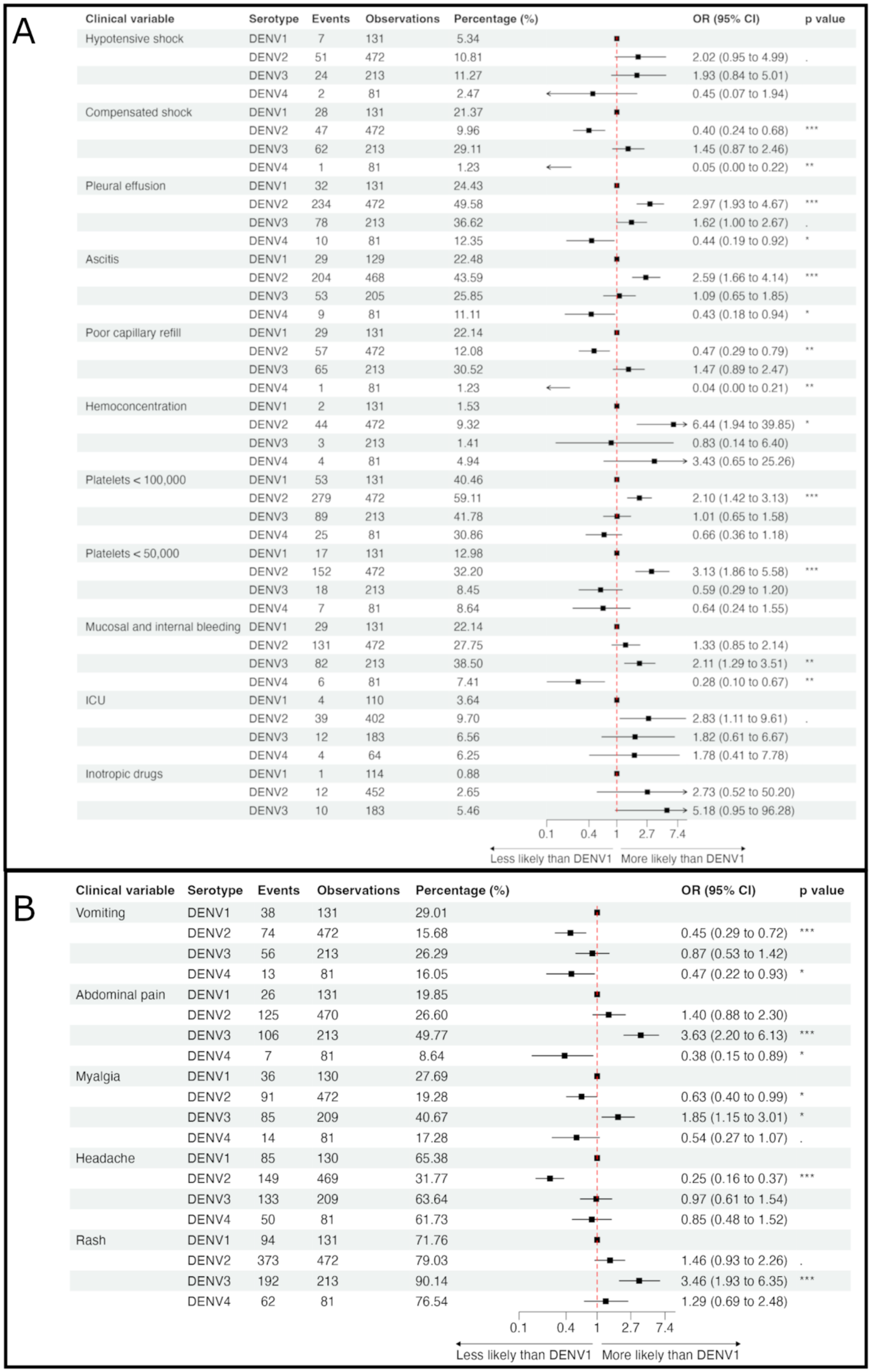


A

| Clinical variable | Serotype | Events | Observations | Percentage (%) | OR (95% CI) | p value |
|---|---|---|---|---|---|---|
| Hypotensive shock | DENV1 | 7 | 131 | 5.34 | | |
| | DENV2 | 51 | 472 | 10.81 | 2.02 (0.95 to 4.99) | . |
| | DENV3 | 24 | 213 | 11.27 | 1.93 (0.84 to 5.01) | |
| | DENV4 | 2 | 81 | 2.47 | 0.45 (0.07 to 1.94) | |
| Compensated shock | DENV1 | 28 | 131 | 21.37 | | |
| | DENV2 | 47 | 472 | 9.96 | 0.40 (0.24 to 0.68) | *** |
| | DENV3 | 62 | 213 | 29.11 | 1.45 (0.87 to 2.46) | |
| | DENV4 | 1 | 81 | 1.23 | 0.05 (0.00 to 0.22) | ** |
| Pleural effusion | DENV1 | 32 | 131 | 24.43 | | |
| | DENV2 | 234 | 472 | 49.58 | 2.97 (1.93 to 4.67) | *** |
| | DENV3 | 78 | 213 | 36.62 | 1.62 (1.00 to 2.67) | . |
| | DENV4 | 10 | 81 | 12.35 | 0.44 (0.19 to 0.92) | * |
| Ascitis | DENV1 | 29 | 129 | 22.48 | | |
| | DENV2 | 204 | 468 | 43.59 | 2.59 (1.66 to 4.14) | *** |
| | DENV3 | 53 | 205 | 25.85 | 1.09 (0.65 to 1.85) | |
| | DENV4 | 9 | 81 | 11.11 | 0.43 (0.18 to 0.94) | * |
| Poor capillary refill | DENV1 | 29 | 131 | 22.14 | | |
| | DENV2 | 57 | 472 | 12.08 | 0.47 (0.29 to 0.79) | ** |
| | DENV3 | 65 | 213 | 30.52 | 1.47 (0.89 to 2.47) | |
| | DENV4 | 1 | 81 | 1.23 | 0.04 (0.00 to 0.21) | ** |
| Hemoconcentration | DENV1 | 2 | 131 | 1.53 | | |
| | DENV2 | 44 | 472 | 9.32 | 6.44 (1.94 to 39.85) | * |
| | DENV3 | 3 | 213 | 1.41 | 0.83 (0.14 to 6.40) | |
| | DENV4 | 4 | 81 | 4.94 | 3.43 (0.65 to 25.26) | |
| Platelets < 100,000 | DENV1 | 53 | 131 | 40.46 | | |
| | DENV2 | 279 | 472 | 59.11 | 2.10 (1.42 to 3.13) | *** |
| | DENV3 | 89 | 213 | 41.78 | 1.01 (0.65 to 1.58) | |
| | DENV4 | 25 | 81 | 30.86 | 0.66 (0.36 to 1.18) | |
| Platelets < 50,000 | DENV1 | 17 | 131 | 12.98 | | |
| | DENV2 | 152 | 472 | 32.20 | 3.13 (1.86 to 5.58) | *** |
| | DENV3 | 18 | 213 | 8.45 | 0.59 (0.29 to 1.20) | |
| | DENV4 | 7 | 81 | 8.64 | 0.64 (0.24 to 1.55) | |
| Mucosal and internal bleeding | DENV1 | 29 | 131 | 22.14 | | |
| | DENV2 | 131 | 472 | 27.75 | 1.33 (0.85 to 2.14) | |
| | DENV3 | 82 | 213 | 38.50 | 2.11 (1.29 to 3.51) | ** |
| | DENV4 | 6 | 81 | 7.41 | 0.28 (0.10 to 0.67) | ** |
| ICU | DENV1 | 4 | 110 | 3.64 | | |
| | DENV2 | 39 | 402 | 9.70 | 2.83 (1.11 to 9.61) | . |
| | DENV3 | 12 | 183 | 6.56 | 1.82 (0.61 to 6.67) | |
| | DENV4 | 4 | 64 | 6.25 | 1.78 (0.41 to 7.78) | |
| Inotropic drugs | DENV1 | 1 | 114 | 0.88 | | |
| | DENV2 | 12 | 452 | 2.65 | 2.73 (0.52 to 50.20) | |
| | DENV3 | 10 | 183 | 5.46 | 5.18 (0.95 to 96.28) | |

B

| Clinical variable | Serotype | Events | Observations | Percentage (%) | OR (95% CI) | p value |
|---|---|---|---|---|---|---|
| Vomiting | DENV1 | 38 | 131 | 29.01 | | |
| | DENV2 | 74 | 472 | 15.68 | 0.45 (0.29 to 0.72) | *** |
| | DENV3 | 56 | 213 | 26.29 | 0.87 (0.53 to 1.42) | |
| | DENV4 | 13 | 81 | 16.05 | 0.47 (0.22 to 0.93) | * |
| Abdominal pain | DENV1 | 26 | 131 | 19.85 | | |
| | DENV2 | 125 | 470 | 26.60 | 1.40 (0.88 to 2.30) | |
| | DENV3 | 106 | 213 | 49.77 | 3.63 (2.20 to 6.13) | *** |
| | DENV4 | 7 | 81 | 8.64 | 0.38 (0.15 to 0.89) | * |
| Myalgia | DENV1 | 36 | 130 | 27.69 | | |
| | DENV2 | 91 | 472 | 19.28 | 0.63 (0.40 to 0.99) | * |
| | DENV3 | 85 | 209 | 40.67 | 1.85 (1.15 to 3.01) | * |
| | DENV4 | 14 | 81 | 17.28 | 0.54 (0.27 to 1.07) | . |
| Headache | DENV1 | 85 | 130 | 65.38 | | |
| | DENV2 | 149 | 469 | 31.77 | 0.25 (0.16 to 0.37) | *** |
| | DENV3 | 133 | 209 | 63.64 | 0.97 (0.61 to 1.54) | |
| | DENV4 | 50 | 81 | 61.73 | 0.85 (0.48 to 1.52) | |
| Rash | DENV1 | 94 | 131 | 71.76 | | |
| | DENV2 | 373 | 472 | 79.03 | 1.46 (0.93 to 2.26) | . |
| | DENV3 | 192 | 213 | 90.14 | 3.46 (1.93 to 6.35) | *** |
| | DENV4 | 62 | 81 | 76.54 | 1.29 (0.69 to 2.48) | |

**Supplementary Figure 7. Clinical signs and symptoms of dengue in secondary cases by serotype in participants of the Pediatric Dengue Hospital-based study, 2004-2022.** A) Clinical signs, laboratory results, and clinical management. B) Symptoms of dengue.

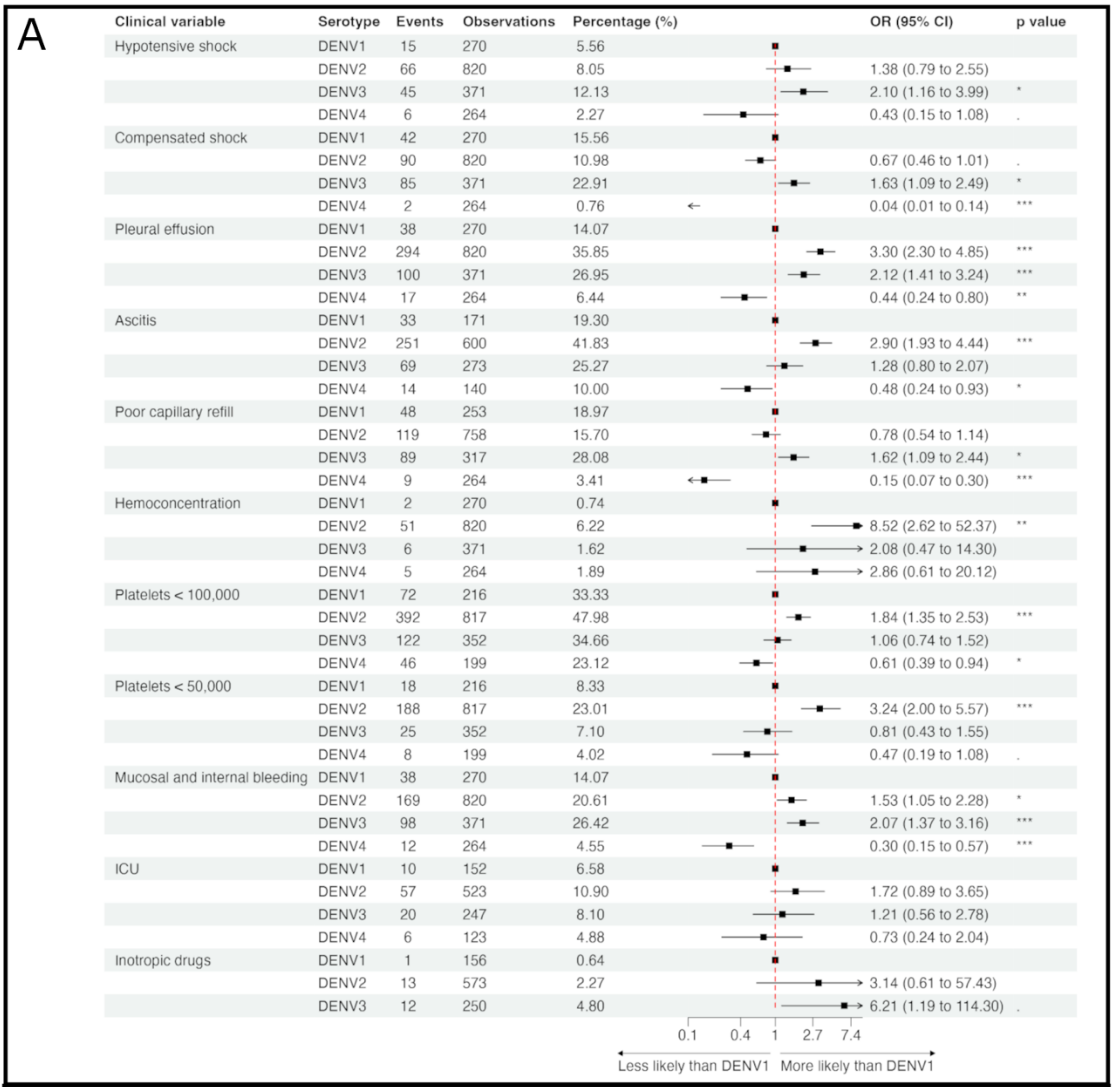

A

| Clinical variable | Serotype | Events | Observations | Percentage (%) | OR (95% CI) | p value |
|---|---|---|---|---|---|---|
| Hypotensive shock | DENV1 | 15 | 270 | 5.56 | | |
| | DENV2 | 66 | 820 | 8.05 | 1.38 (0.79 to 2.55) | |
| | DENV3 | 45 | 371 | 12.13 | 2.10 (1.16 to 3.99) | * |
| | DENV4 | 6 | 264 | 2.27 | 0.43 (0.15 to 1.08) | . |
| Compensated shock | DENV1 | 42 | 270 | 15.56 | | |
| | DENV2 | 90 | 820 | 10.98 | 0.67 (0.46 to 1.01) | . |
| | DENV3 | 85 | 371 | 22.91 | 1.63 (1.09 to 2.49) | * |
| | DENV4 | 2 | 264 | 0.76 | 0.04 (0.01 to 0.14) | *** |
| Pleural effusion | DENV1 | 38 | 270 | 14.07 | | |
| | DENV2 | 294 | 820 | 35.85 | 3.30 (2.30 to 4.85) | *** |
| | DENV3 | 100 | 371 | 26.95 | 2.12 (1.41 to 3.24) | *** |
| | DENV4 | 17 | 264 | 6.44 | 0.44 (0.24 to 0.80) | ** |
| Ascitis | DENV1 | 33 | 171 | 19.30 | | |
| | DENV2 | 251 | 600 | 41.83 | 2.90 (1.93 to 4.44) | *** |
| | DENV3 | 69 | 273 | 25.27 | 1.28 (0.80 to 2.07) | |
| | DENV4 | 14 | 140 | 10.00 | 0.48 (0.24 to 0.93) | * |
| Poor capillary refill | DENV1 | 48 | 253 | 18.97 | | |
| | DENV2 | 119 | 758 | 15.70 | 0.78 (0.54 to 1.14) | |
| | DENV3 | 89 | 317 | 28.08 | 1.62 (1.09 to 2.44) | * |
| | DENV4 | 9 | 264 | 3.41 | 0.15 (0.07 to 0.30) | *** |
| Hemoconcentration | DENV1 | 2 | 270 | 0.74 | | |
| | DENV2 | 51 | 820 | 6.22 | 8.52 (2.62 to 52.37) | ** |
| | DENV3 | 6 | 371 | 1.62 | 2.08 (0.47 to 14.30) | |
| | DENV4 | 5 | 264 | 1.89 | 2.86 (0.61 to 20.12) | |
| Platelets < 100,000 | DENV1 | 72 | 216 | 33.33 | | |
| | DENV2 | 392 | 817 | 47.98 | 1.84 (1.35 to 2.53) | *** |
| | DENV3 | 122 | 352 | 34.66 | 1.06 (0.74 to 1.52) | |
| | DENV4 | 46 | 199 | 23.12 | 0.61 (0.39 to 0.94) | * |
| Platelets < 50,000 | DENV1 | 18 | 216 | 8.33 | | |
| | DENV2 | 188 | 817 | 23.01 | 3.24 (2.00 to 5.57) | *** |
| | DENV3 | 25 | 352 | 7.10 | 0.81 (0.43 to 1.55) | |
| | DENV4 | 8 | 199 | 4.02 | 0.47 (0.19 to 1.08) | . |
| Mucosal and internal bleeding | DENV1 | 38 | 270 | 14.07 | | |
| | DENV2 | 169 | 820 | 20.61 | 1.53 (1.05 to 2.28) | * |
| | DENV3 | 98 | 371 | 26.42 | 2.07 (1.37 to 3.16) | *** |
| | DENV4 | 12 | 264 | 4.55 | 0.30 (0.15 to 0.57) | *** |
| ICU | DENV1 | 10 | 152 | 6.58 | | |
| | DENV2 | 57 | 523 | 10.90 | 1.72 (0.89 to 3.65) | |
| | DENV3 | 20 | 247 | 8.10 | 1.21 (0.56 to 2.78) | |
| | DENV4 | 6 | 123 | 4.88 | 0.73 (0.24 to 2.04) | |
| Inotropic drugs | DENV1 | 1 | 156 | 0.64 | | |
| | DENV2 | 13 | 573 | 2.27 | 3.14 (0.61 to 57.43) | |
| | DENV3 | 12 | 250 | 4.80 | 6.21 (1.19 to 114.30) | . |



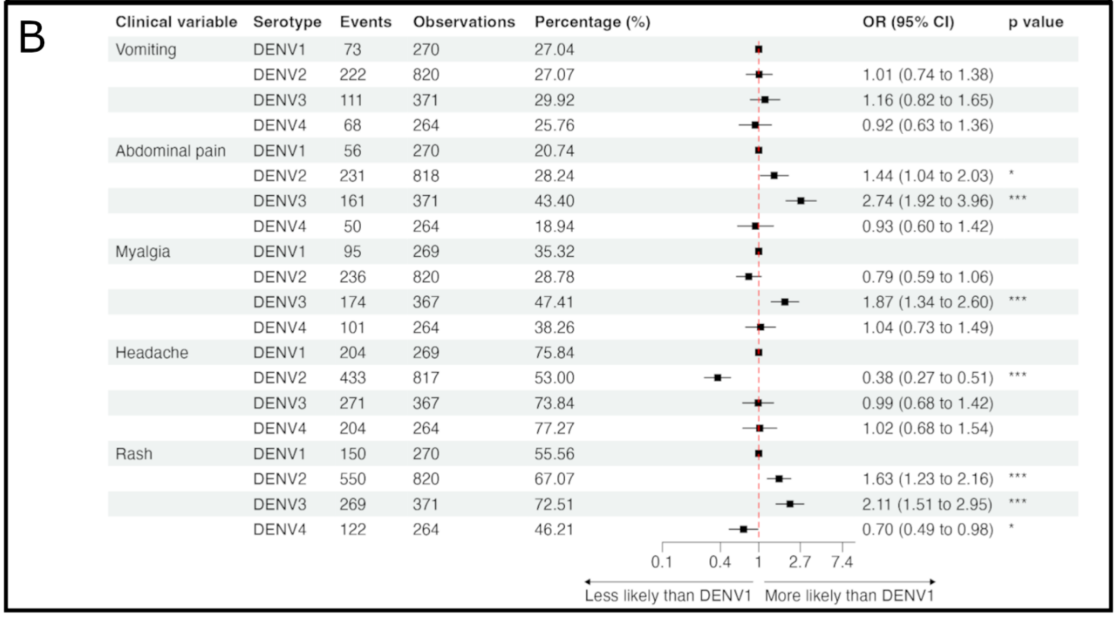

B

| Clinical variable | Serotype | Events | Observations | Percentage (%) | OR (95% CI) | p value |
|---|---|---|---|---|---|---|
| Vomiting | DENV1 | 73 | 270 | 27.04 | | |
| | DENV2 | 222 | 820 | 27.07 | 1.01 (0.74 to 1.38) | |
| | DENV3 | 111 | 371 | 29.92 | 1.16 (0.82 to 1.65) | |
| | DENV4 | 68 | 264 | 25.76 | 0.92 (0.63 to 1.36) | |
| Abdominal pain | DENV1 | 56 | 270 | 20.74 | | |
| | DENV2 | 231 | 818 | 28.24 | 1.44 (1.04 to 2.03) | * |
| | DENV3 | 161 | 371 | 43.40 | 2.74 (1.92 to 3.96) | *** |
| | DENV4 | 50 | 264 | 18.94 | 0.93 (0.60 to 1.42) | |
| Myalgia | DENV1 | 95 | 269 | 35.32 | | |
| | DENV2 | 236 | 820 | 28.78 | 0.79 (0.59 to 1.06) | |
| | DENV3 | 174 | 367 | 47.41 | 1.87 (1.34 to 2.60) | *** |
| | DENV4 | 101 | 264 | 38.26 | 1.04 (0.73 to 1.49) | |
| Headache | DENV1 | 204 | 269 | 75.84 | | |
| | DENV2 | 433 | 817 | 53.00 | 0.38 (0.27 to 0.51) | *** |
| | DENV3 | 271 | 367 | 73.84 | 0.99 (0.68 to 1.42) | |
| | DENV4 | 204 | 264 | 77.27 | 1.02 (0.68 to 1.54) | |
| Rash | DENV1 | 150 | 270 | 55.56 | | |
| | DENV2 | 550 | 820 | 67.07 | 1.63 (1.23 to 2.16) | *** |
| | DENV3 | 269 | 371 | 72.51 | 2.11 (1.51 to 2.95) | *** |
| | DENV4 | 122 | 264 | 46.21 | 0.70 (0.49 to 0.98) | * |



**Supplementary Figure 8. Clinical signs and symptoms of dengue in secondary cases by serotype in participants of the Pediatric Dengue Cohort Study transferred to the Pediatric Dengue Hospital-based study.** A) Clinical signs, laboratory results, and clinical management. B) Symptoms of dengue.

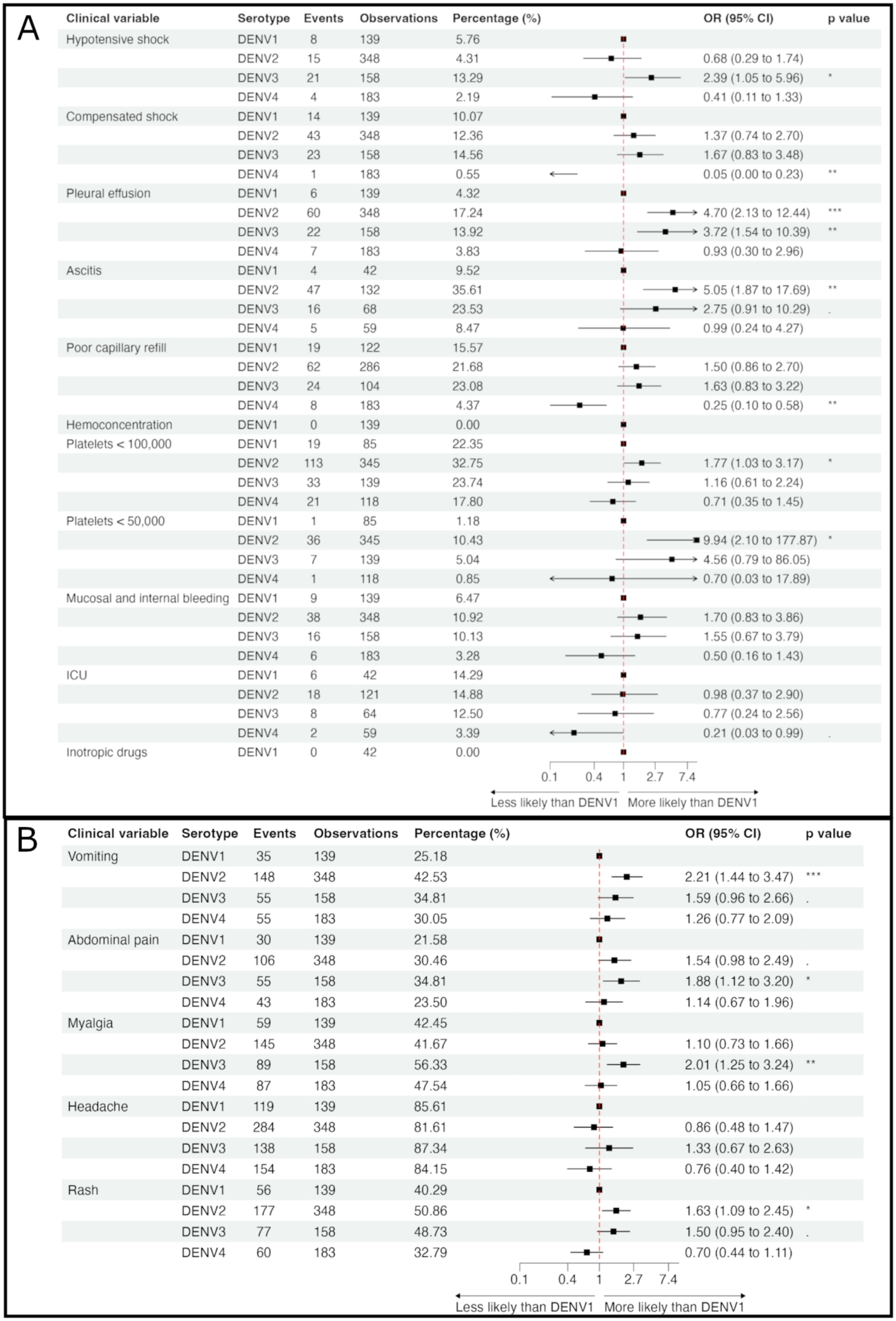

| Clinical variable | Serotype | Events | Observations | Percentage (%) | OR (95% CI) | p value |
|---|---|---|---|---|---|---|
| Hypotensive shock | DENV1 | 8 | 139 | 5.76 | | |
| | DENV2 | 15 | 348 | 4.31 | 0.68 (0.29 to 1.74) | |
| | DENV3 | 21 | 158 | 13.29 | 2.39 (1.05 to 5.96) | * |
| | DENV4 | 4 | 183 | 2.19 | 0.41 (0.11 to 1.33) | |
| Compensated shock | DENV1 | 14 | 139 | 10.07 | | |
| | DENV2 | 43 | 348 | 12.36 | 1.37 (0.74 to 2.70) | |
| | DENV3 | 23 | 158 | 14.56 | 1.67 (0.83 to 3.48) | |
| | DENV4 | 1 | 183 | 0.55 | 0.05 (0.00 to 0.23) | ** |
| Pleural effusion | DENV1 | 6 | 139 | 4.32 | | |
| | DENV2 | 60 | 348 | 17.24 | 4.70 (2.13 to 12.44) | *** |
| | DENV3 | 22 | 158 | 13.92 | 3.72 (1.54 to 10.39) | ** |
| | DENV4 | 7 | 183 | 3.83 | 0.93 (0.30 to 2.96) | |
| Ascitis | DENV1 | 4 | 42 | 9.52 | | |
| | DENV2 | 47 | 132 | 35.61 | 5.05 (1.87 to 17.69) | ** |
| | DENV3 | 16 | 68 | 23.53 | 2.75 (0.91 to 10.29) | . |
| | DENV4 | 5 | 59 | 8.47 | 0.99 (0.24 to 4.27) | |
| Poor capillary refill | DENV1 | 19 | 122 | 15.57 | | |
| | DENV2 | 62 | 286 | 21.68 | 1.50 (0.86 to 2.70) | |
| | DENV3 | 24 | 104 | 23.08 | 1.63 (0.83 to 3.22) | |
| | DENV4 | 8 | 183 | 4.37 | 0.25 (0.10 to 0.58) | ** |
| Hemoconcentration | DENV1 | 0 | 139 | 0.00 | | |
| Platelets < 100,000 | DENV1 | 19 | 85 | 22.35 | | |
| | DENV2 | 113 | 345 | 32.75 | 1.77 (1.03 to 3.17) | * |
| | DENV3 | 33 | 139 | 23.74 | 1.16 (0.61 to 2.24) | |
| | DENV4 | 21 | 118 | 17.80 | 0.71 (0.35 to 1.45) | |
| Platelets < 50,000 | DENV1 | 1 | 85 | 1.18 | | |
| | DENV2 | 36 | 345 | 10.43 | 9.94 (2.10 to 177.87) | * |
| | DENV3 | 7 | 139 | 5.04 | 4.56 (0.79 to 86.05) | |
| | DENV4 | 1 | 118 | 0.85 | 0.70 (0.03 to 17.89) | |
| Mucosal and internal bleeding | DENV1 | 9 | 139 | 6.47 | | |
| | DENV2 | 38 | 348 | 10.92 | 1.70 (0.83 to 3.86) | |
| | DENV3 | 16 | 158 | 10.13 | 1.55 (0.67 to 3.79) | |
| | DENV4 | 6 | 183 | 3.28 | 0.50 (0.16 to 1.43) | |
| ICU | DENV1 | 6 | 42 | 14.29 | | |
| | DENV2 | 18 | 121 | 14.88 | 0.98 (0.37 to 2.90) | |
| | DENV3 | 8 | 64 | 12.50 | 0.77 (0.24 to 2.56) | |
| | DENV4 | 2 | 59 | 3.39 | 0.21 (0.03 to 0.99) | . |
| Inotropic drugs | DENV1 | 0 | 42 | 0.00 | | |



| Clinical variable | Serotype | Events | Observations | Percentage (%) | OR (95% CI) | p value |
|---|---|---|---|---|---|---|
| Vomiting | DENV1 | 35 | 139 | 25.18 | | |
| | DENV2 | 148 | 348 | 42.53 | 2.21 (1.44 to 3.47) | *** |
| | DENV3 | 55 | 158 | 34.81 | 1.59 (0.96 to 2.66) | . |
| | DENV4 | 55 | 183 | 30.05 | 1.26 (0.77 to 2.09) | |
| Abdominal pain | DENV1 | 30 | 139 | 21.58 | | |
| | DENV2 | 106 | 348 | 30.46 | 1.54 (0.98 to 2.49) | . |
| | DENV3 | 55 | 158 | 34.81 | 1.88 (1.12 to 3.20) | * |
| | DENV4 | 43 | 183 | 23.50 | 1.14 (0.67 to 1.96) | |
| Myalgia | DENV1 | 59 | 139 | 42.45 | | |
| | DENV2 | 145 | 348 | 41.67 | 1.10 (0.73 to 1.66) | |
| | DENV3 | 89 | 158 | 56.33 | 2.01 (1.25 to 3.24) | ** |
| | DENV4 | 87 | 183 | 47.54 | 1.05 (0.66 to 1.66) | |
| Headache | DENV1 | 119 | 139 | 85.61 | | |
| | DENV2 | 284 | 348 | 81.61 | 0.86 (0.48 to 1.47) | |
| | DENV3 | 138 | 158 | 87.34 | 1.33 (0.67 to 2.63) | |
| | DENV4 | 154 | 183 | 84.15 | 0.76 (0.40 to 1.42) | |
| Rash | DENV1 | 56 | 139 | 40.29 | | |
| | DENV2 | 177 | 348 | 50.86 | 1.63 (1.09 to 2.45) | * |
| | DENV3 | 77 | 158 | 48.73 | 1.50 (0.95 to 2.40) | . |
| | DENV4 | 60 | 183 | 32.79 | 0.70 (0.44 to 1.11) | |



**Supplemental Figure 9. Clinical signs and symptoms of dengue in secondary cases by serotype in participants of the Pediatric Dengue Cohort Study, 2004-2022.** A) Clinical signs, laboratory results, and clinical management. B) Symptoms of dengue.
